# Decision Analysis Modeling Favors Optimal Influenza Vaccination in Late November or Early December

**DOI:** 10.64898/2026.09.11.26362843

**Authors:** Austin G. Meyer, Brittany N. Rosales, Shihao Yang, Mauricio Santillana

**Affiliations:** Division of Pediatric Hospital Medicine, Department of Pediatrics, Baylor Scott and White Health, Temple, TX, USA; Department of Pediatrics, Baylor College of Medicine, Temple, TX, USA; Division of Pulmonary, Critical Care and Sleep Medicine, Dell Medical School, University of Texas at Austin, Austin, TX 78712, USA; H. Milton Stewart School of Industrial and Systems Engineering, Georgia Institute of Technology, Atlanta, GA, USA; Machine Intelligence Group for the Betterment of Health and the Environment (MIGHTE) Lab, Department of Physics, Northeastern University, Boston, MA, USA

**Author notes:** **Corresponding author:** Austin G. Meyer, MD, PhD, MS, MPH, MS; Division of Pediatric Hospital Medicine, Department of Pediatrics, Baylor Scott and White Health; 1901 SW H K Dodgen Loop, MS-CH-1139, Temple, TX 76502, USA. **Financial support:** None.

## Abstract

**Background:** Waning vaccine protection and variable epidemic timing create a tradeoff between vaccinating before influenza circulation and preserving protection when disease burden is greatest. Existing guidance does not quantify the week that best balances this tradeoff.

**Objective:** To estimate the optimal week for influenza vaccination while accounting for uncertainty in epidemic timing and vaccine protection.

**Design:** Simulation-based probabilistic decision analysis.

**Setting:** United States outpatient influenza surveillance from the 50 states and District of Columbia during the 2010/11 through 2018/19 and 2023/24 through 2025/26 seasons.

**Participants:** People in the United States who would receive 1 influenza vaccine dose.

**Interventions:** Vaccination during Morbidity and Mortality Weekly Report (MMWR) weeks 36 through 12.

**Measurements:** We combined influenza-like illness (ILI) syndromic surveillance and influenza-specific data with uncertainty in epidemic timing, initial vaccine effectiveness, waning, and immune-response lag. For each candidate week, the model estimated direct protection against outpatient surveillance burden.

**Results:** The primary model identified the optimal time to vaccinate as week 47, with weeks 47–48 being of similar utility. Our influenza-weighted outpatient analysis (ILI+) selected week 48. Compared with week 44, near the end of the current September–October guidance window, week 48 reduced modeled influenza-weighted outpatient burden by 50 visit equivalents per 10,000 weekly encounters.

**Limitations:** The model assumed vaccination would occur, used outpatient surveillance burden rather than severe outcomes, and did not incorporate transmission effects.

**Conclusion:** Under the primary waning model, these findings favor vaccinating during weeks 47–48, approximately late November through early December, for most one-dose recipients.

**Primary Funding Source:** None.

## Introduction

Seasonal influenza continues to cause substantial annual morbidity and mortality in the United States (US) (1). Vaccination is our principal means of preventing this impact (2, 3). The Advisory Committee on Immunization Practices (ACIP) recommends routine annual influenza vaccination for everyone at least 6 months of age who has no contraindication. For most people, the guidance recommends one dose of vaccine during September or October, and that vaccination continue to be offered for those not yet vaccinated while influenza continues to circulate. In addition, current guidance explicitly discourages July and August vaccination for most groups because accumulating evidence suggests that vaccine-induced protection may wane during the season (4).

Over the last two decades, several US and international studies have found lower vaccine effectiveness (VE) with increasing time since vaccination with the size of this association varying by season, virus subtype, age group, and study design (5–8). While analysis of the waning effect is challenging due to selection bias, depletion of susceptible patients, and other estimation challenges (9–11), good evidence of any waning effect creates a timing tradeoff between vaccination and the annual epidemic. If a person is vaccinated earlier, their vaccine-induced immunity may diminish substantially over time and leave them at higher risk during a significant portion of the season. By contrast, if a person is vaccinated late, they may be at risk during the start of the season as their immunity builds or if the season peaks earlier than expected in their area. Clinicians and patients therefore face a practical question: when should a person who plans to be vaccinated receive the vaccine?

Previous studies have evaluated this tradeoff to some extent with several approaches including economic models, historical uptake schedules, and counterfactual shifts in vaccination timing. These analyses have variously compared monthly schedules for older US adults, vaccine campaign start dates for US older adults, monthly timing using Australian age-specific notification data, and changes to observed schedules for US older adults (12–15). Most recently, Spencer and colleagues shifted observed vaccine uptake schedules for five age groups across ten US seasons under six combinations of initial VE and waning (16).

These studies help to confirm that ideal vaccination timing depends on epidemic timing, waning, and initial protection. They generally test several possible scenarios and report scenario-specific optima or changes in outcome burden (infection, hospitalization, etc.) under shifted vaccination schedules. While these studies are scientifically valuable, collectively, they lack simple applicability for most people in the US. This issue stems from the fact that several studies are not generalizable to the US population; some focus on public health vaccine campaign start times rather than providing a person-level recommendation to patients and clinicians; and others provide optimal timing under conditions that generally cannot be known to patients or clinicians before the season. For example, while Spencer et al. show that the best vaccination schedule is sensitive to VE, waning rate, and peak timing, vaccine effectiveness and peak timing cannot be estimated sufficiently far in advance to incorporate this information into clinical decision-making. In addition, the incorporation of vaccine uptake curves is helpful for public health officials designing vaccination campaigns, but reduces utility for individual patient-level decision-making. Moreover, with an alternative modeling framework it may not be necessary to have accurate live forecasts to make an optimal or near-optimal vaccine timing decision for an individual. For that reason, the available studies do not provide an actionable, quantitative estimate of the optimal week that integrates current evidence and its uncertainty for individual patient decision-making.

We developed a simulation-based probabilistic decision model to estimate optimal vaccination timing for a person who plans to receive one influenza vaccine dose. The model combined smoothed historical influenza-like illness (ILI) curves and influenza-specific curves with uncertainty in epidemic timing, curve estimation, baseline VE, waning, and immune-response lag. We aimed to identify a fixed week that could be chosen before the season and to estimate how much modeled protection would be forgone by vaccinating earlier or later.

## Methods

### Study Design and Data

We conducted a simulation-based decision analysis to choose a vaccination week for a person who will receive one influenza vaccine dose. Candidate dates were Morbidity and Mortality Weekly Report (MMWR) weeks 36–52 and 1–12, with protection evaluated during weeks 36–22 of the influenza season.

The primary outcome curve was the weekly proportion of outpatient visits for ILI reported to the U.S. Outpatient Influenza-like Illness Surveillance Network (ILINet) of the Centers for Disease Control and Prevention (CDC) (17). We restricted the analysis to the 50 states and the District of Columbia. We began with 2010/11, the first season after the 2009 H1N1 pandemic and the first under universal U.S. influenza vaccination guidance (18). We included seasons through 2018/19 and 2023/24 through 2025/26, excluding those most disrupted by the coronavirus disease 2019 (COVID-19) pandemic. We combined state curves using population weights and allocated modeled ILI burden among 5 age groups using the weekly age distribution of ILI visits in each Department of Health and Human Services (HHS) region. National laboratory influenza positivity supported secondary analyses. Full definitions are provided in the Supplement.

### Decision Model

We fit smooth seasonal ILI curves separately for each state. Each of 5,000 simulations selected a historical season, sampled a curve from the fitted model, and allowed a small shift in epidemic timing. The same season and shift were used in every state, preserving geographic timing patterns.

Each simulation also sampled vaccine effectiveness independently of the epidemic season. All-age analyses used overall estimates from 2010/11 through 2018/19 and 2023/24 through 2025/26, with a preliminary estimate for 2025/26. Age-specific analyses used age-matched estimates from 2010/11 through 2018/19 and 2023/24 (19–23). Supplement Tables S1–S2 list VE estimates and other empirical inputs with their sources. Supplement Figures S3–S4 show the seasonal burden and vaccine-protection distributions. Full protection developed 1, 2, or 3 weeks after vaccination, centered on 2 weeks, and then waned according to the estimate from Ray and colleagues (6, 18).

We treated each source effectiveness estimate as applying 8 weeks after full protection developed and used the same waning distribution across ages and seasons.

For each possible vaccination week, we multiplied the protection remaining each week by that week’s sampled outpatient burden and summed across the season. Regret was the protection that was not achieved (i.e., counterfactual protection) relative to the best week in that simulation. We chose the fixed week with the lowest mean regret, which is also the week with the greatest mean protected burden. A near-optimal week retained at least 95% of the protection available from the best week. We expressed regret as standardized ILI visits over the season in a system with 10,000 outpatient encounters each week.

### Sensitivity Analyses

We repeated the analysis using an unsmoothed empirical ILINet bootstrap and, separately, using only pre-pandemic burden seasons. In addition, we used alternative curve flexibility, immune-response times, waning rates, and vaccine-effectiveness reference times. Other analyses used laboratory positivity alone or multiplied by ILI, and omitted each season in turn for evaluation. Each analysis used 5,000 simulations. Analyses were conducted in R version 4.5.1 (24). Full details are provided in the Supplement. Code to retrieve the public data and run the complete reproducibility workflow will be available at https://github.com/ausmeyer/flu_optimal_vaccine_estimate.

### Ethics

All analyses used publicly available, aggregate surveillance and population data. Because no individual-level or identifiable information was used, this study did not constitute human subjects research and did not require institutional review board review.

### Role of the Funding Source

This work received no specific funding. No funding source had a role in the study design; data collection, analysis, or interpretation; manuscript preparation; or the decision to submit the manuscript for publication.

## Results

### Vaccination Timing by Age Group and State

The 12 included influenza seasons differed substantially in timing (Figure 1). Time was measured in MMWR weeks, which are CDC epidemiologic weeks whose calendar dates shift slightly from year to year. National ILI peaks ranged from MMWR week 52 to week 11 such that no single typical epidemic curve could represent the timing uncertainty in this decision. We therefore sampled across historical seasons when choosing a vaccination week. For reference, weeks 43–44 typically fall near the end of October, week 47 typically falls in mid-to-late November, and week 48 can extend into early December.

**Figure 1.**
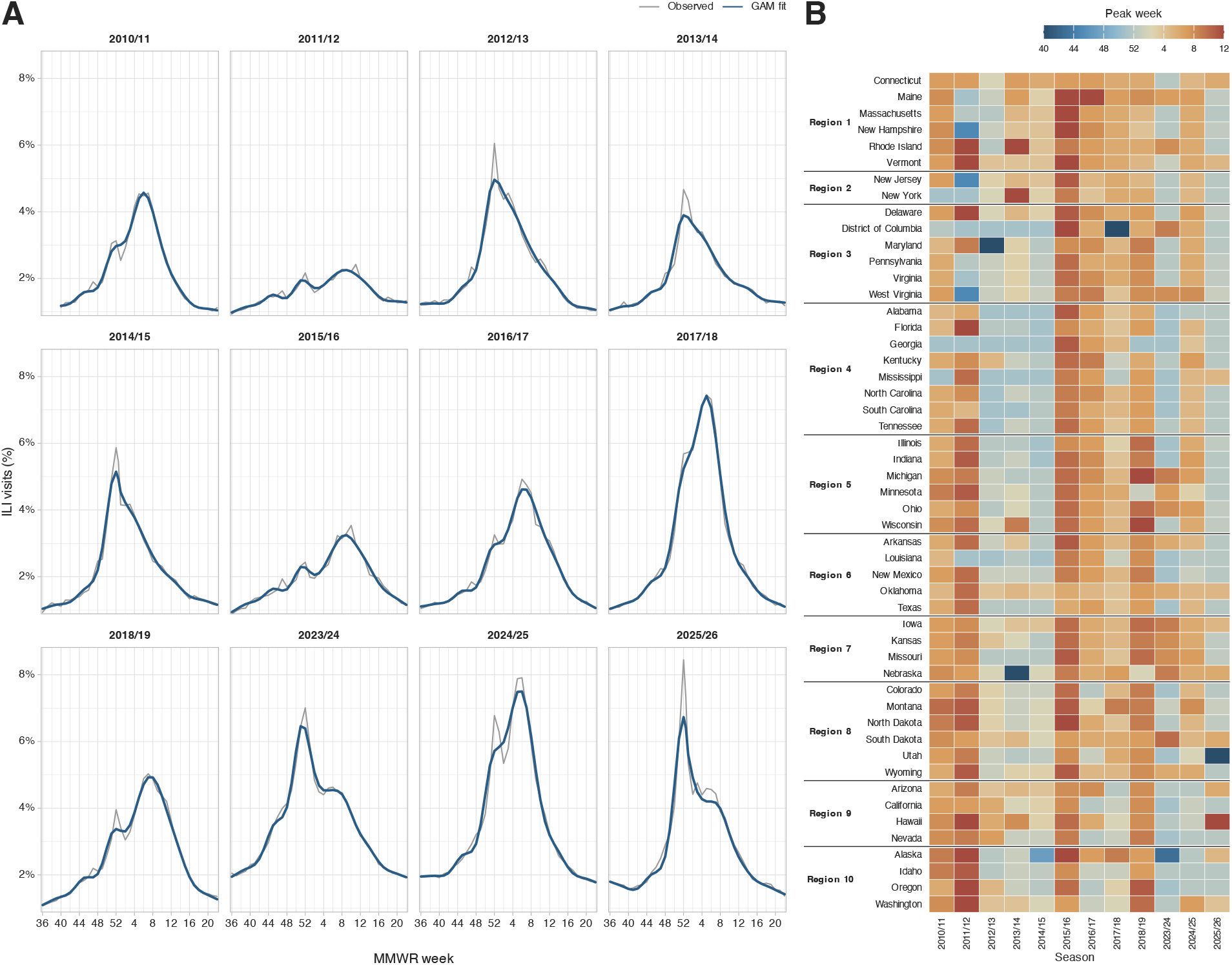
Historical influenza epidemic timing. (A) Observed and GAM-fitted national ILI curves for each of the 12 included seasons. Gray lines show the observed population-weighted ILI proportion; blue lines show the GAM-fitted national curve over available weeks. The 2010/11 curves begin at MMWR week 40; all others begin at week 36, and all extend through week 22. (B) GAM-fitted peak week for each state and season. States are grouped by HHS region. Color shows earlier peaks in blue and later peaks in red.

We measured timing quality as regret: the modeled avoidable outpatient disease burden in a particular week relative to vaccination in the best possible week in each simulation. We combined many individual simulation draws to characterize the distribution of regret. For the all-age population, MMWR week 47 minimized mean regret and was the primary optimal time to vaccinate (Figure 2). Week 48 performed nearly as well, producing a broad trough across weeks 47–48. The median best week across simulations was week 48, with half of draws having an optimum falling between weeks 47 and 49. In a surveillance system with 10,000 outpatient encounters each week, mean regret was 189 visits for vaccination in week 44, 55 in week 47, 55 in week 48, and 380 in week 52. Regret remains above zero at the optimal week because no one fixed week is best in every simulated season.

**Figure 2.**
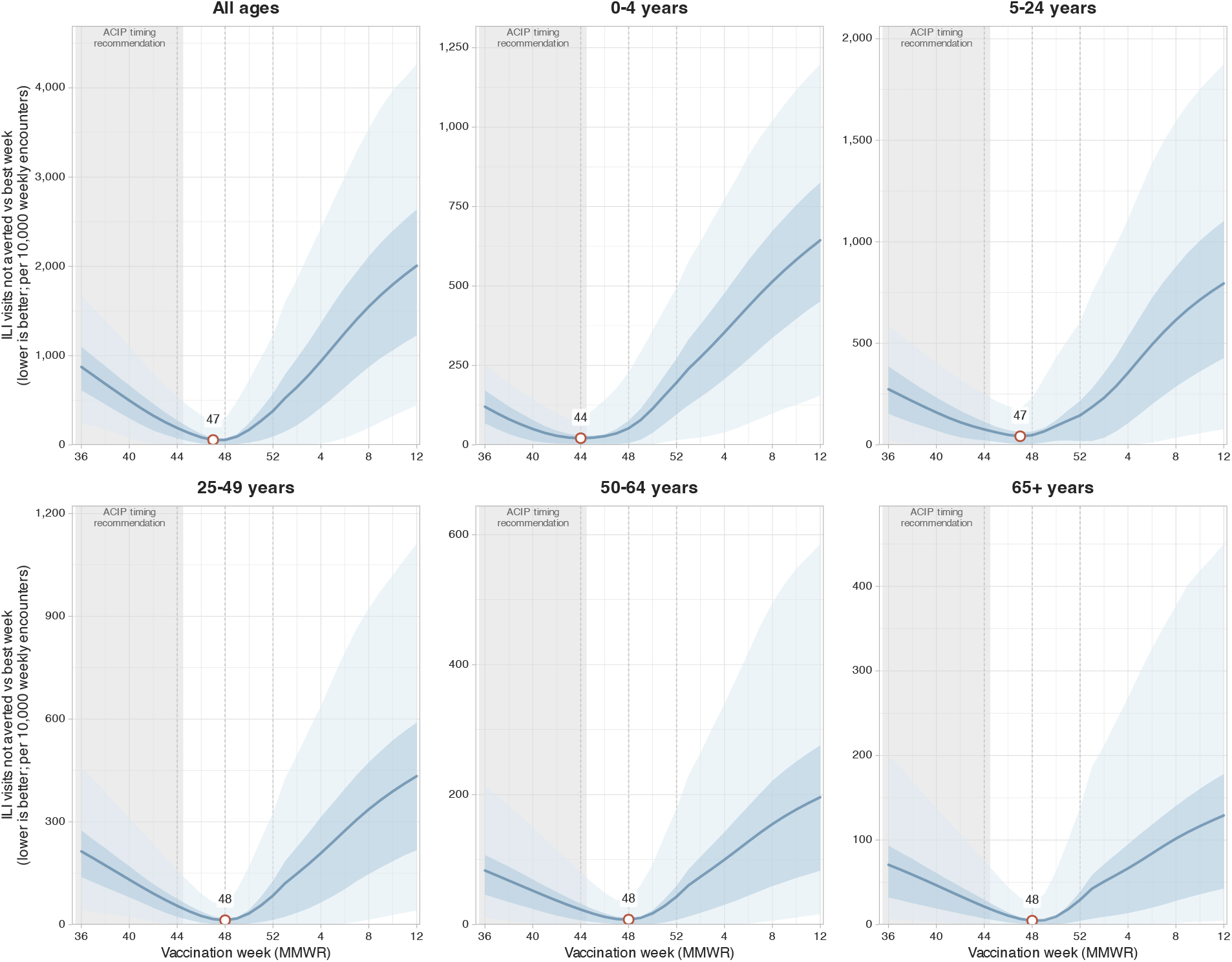
National expected regret by vaccination week and age group. Abbreviations: ACIP, Advisory Committee on Immunization Practices; ILI, influenza-like illness; MMWR, Morbidity and Mortality Weekly Report; vs, versus. Regret is the modeled ILI burden left avoidable relative to the best week in the same simulation, shown as standardized ILI visits over the season in a system with 10,000 outpatient encounters each week. Lower values indicate better timing. Solid lines show mean regret; bands show the central 50% and 95% of simulation-specific regret. Open points mark the week that minimized mean regret. Dashed lines mark weeks 44, 48, and 52. The shaded region approximates the ACIP September–October guidance window. Vertical scales differ across panels, so compare panels by timing rather than magnitude.

For each specific age group, the regret-minimizing week was 44 for ages 0–4 years, compared with week 47 for ages 5–24 years and week 48 for ages 25–49, 50–64, and 65 years or older (Figure 2). Since the analysis evaluates one dose, the age-specific results do not apply to any child who requires two doses that season, including many of the 0–4-year-olds and some 5–8-year-olds. In addition, the week-44 result also does not apply to infants younger than 6 months, who contribute to the ILI surveillance outcome but are not eligible for influenza vaccination.

State-level variation was modest for the all-age timing decision as well as for older age groups (Figure 3A). Regret-minimizing weeks ranged from 46 to 50 across states, and 45 of 51 locations selected weeks 47–49. The ranges were wider for ages 0–4 years (weeks 41–48) and ages 5–24 years (weeks 44–51), but stayed within weeks 46–50 for each of the three older age groups. In addition, earlier optimal timing for ages 0–4 years was concentrated in Department of Health and Human Services (HHS) Regions 2–4.

**Figure 3.**
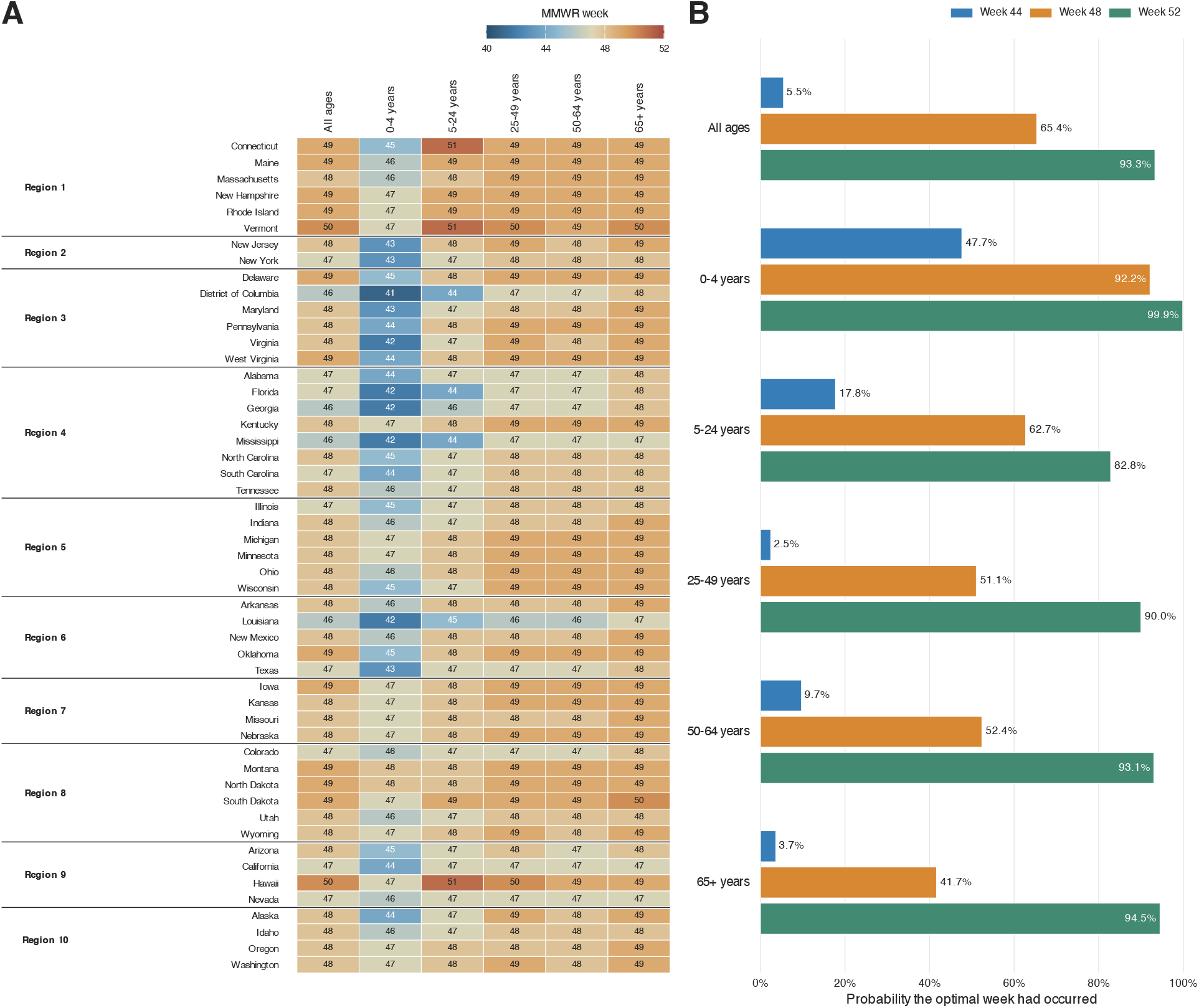
State and age-specific vaccination timing and when the best week had occurred. (A) Week that minimized mean regret for each state and age group, including the all-age population. States are grouped by HHS region. The color scale spans weeks 40–52, with earlier timing in blue and later timing in red. (B) National probabilities for the United States, by age group, that the best week in each simulation had already occurred by MMWR weeks 44, 48, and 52. These are not probabilities that vaccination by that week is effective.

### Consequences of Vaccination During Common Calendar Windows

A week can be best in relatively few simulations and still perform well in many of them. For all ages, week 47 retained at least 95% of the protection obtained by vaccinating in a particular simulation’s best week in 83.1% of simulations, and week 48 did so in 86.0% of simulations. Likewise, week 44 retained 95% of the protection of the best week in 40.8% of simulations, and week 40 in only 8.1% of simulations.

In terms of direct clinical consequence, ACIP recommends influenza vaccination during September or October for most people (4). Week 44 generally falls at the end of that window or slightly past it and week 40 falls in the middle of the recommended range. Relative to the best timing in the same simulation, vaccination in week 40 lost an average of 21.8% of achievable modeled protection against outpatient ILI surveillance burden, and week 44 lost 9.2%. By contrast, the protection losses were 2.8% in week 47 and 2.3% in week 48.

For all ages, the best week in each simulation usually occurred after the September–October window, but the pattern differed for children aged 0–4 years. To better understand this difference we computed the chance that the best vaccination week had already occurred by a given date cutpoint (Figure 3B). For the all-age population, that probability was only 5.5% by week 44 (approximately the end of October), 65.4% by week 48, and 93.3% by week 52. For ages 0–4 years, the best week had occurred by week 44 in 47.7% of simulations, compared with 2.5% to 17.8% for older age groups. By week 48, the corresponding probabilities were 92.2% for ages 0–4 years and 41.7% to 62.7% for older groups (Figure 3B).

### Robustness Across Assumptions and Seasons

Fourteen of the 17 sensitivity analyses selected weeks 47–48. The three analyses using alternative cubic waning curves selected weeks 40–46 (Figure 4; Table 1).

**Table 1.** Primary and sensitivity analyses. For each analysis, the table reports the week that minimized mean regret and the chance that the best week in each simulation had occurred by MMWR weeks 44, 48, and 52. The selected week is the single decision estimate. Probabilities are shown to one decimal place. Values below 0.1% and above 99.9% are shown as thresholds because they rest on one or two of the 5,000 simulations.

| Analysis | Selected week | By week 44 | By week 48 | By week 52 |
| --- | --- | --- | --- | --- |
| Primary GAM | 47 | 5.5% | 65.4% | 93.3% |
| No epidemic timing shift | 48 | 5.5% | 67.7% | 93.1% |
| Unsmoothed empirical ILINet | 48 | 7.7% | 58.3% | 92.1% |
| ILI+, population-weighted ILI | 48 | 3.6% | 52.0% | 81.2% |
| ILI+, unweighted ILI | 48 | 3.8% | 52.4% | 81.2% |
| ILI+, clinical laboratories only | 48 | 1.8% | 46.6% | 93.2% |
| Pre-pandemic burden GAM | 48 | 4.6% | 58.8% | 90.3% |
| Fixed 2-week immune lag | 48 | 5.3% | 66.4% | 93.3% |
| Ray slow waning | 47 | 12.7% | 74.1% | 95.8% |
| Ray fast waning | 48 | 1.3% | 55.6% | 90.3% |
| Spencer–Ferdinands fast waning | 46 | 13.3% | 99.0% | >99.9% |
| Spencer–Ferdinands, onset aligned | 45 | 20.4% | 99.6% | >99.9% |
| Spencer slow waning | 40 | >99.9% | >99.9% | >99.9% |
| Higher spline basis | 48 | 7.0% | 61.1% | 93.6% |
| VE reference at 4 weeks | 48 | 1.8% | 49.5% | 85.4% |
| VE reference at 12 weeks | 47 | 15.3% | 80.4% | 98.2% |
| ICLS/NREVSS national | 48 | 2.4% | 50.7% | 78.6% |
| ICLS/NREVSS influenza A, 2015/16 onward | 48 | 0.8% | 48.6% | 82.3% |

**Figure 4.**
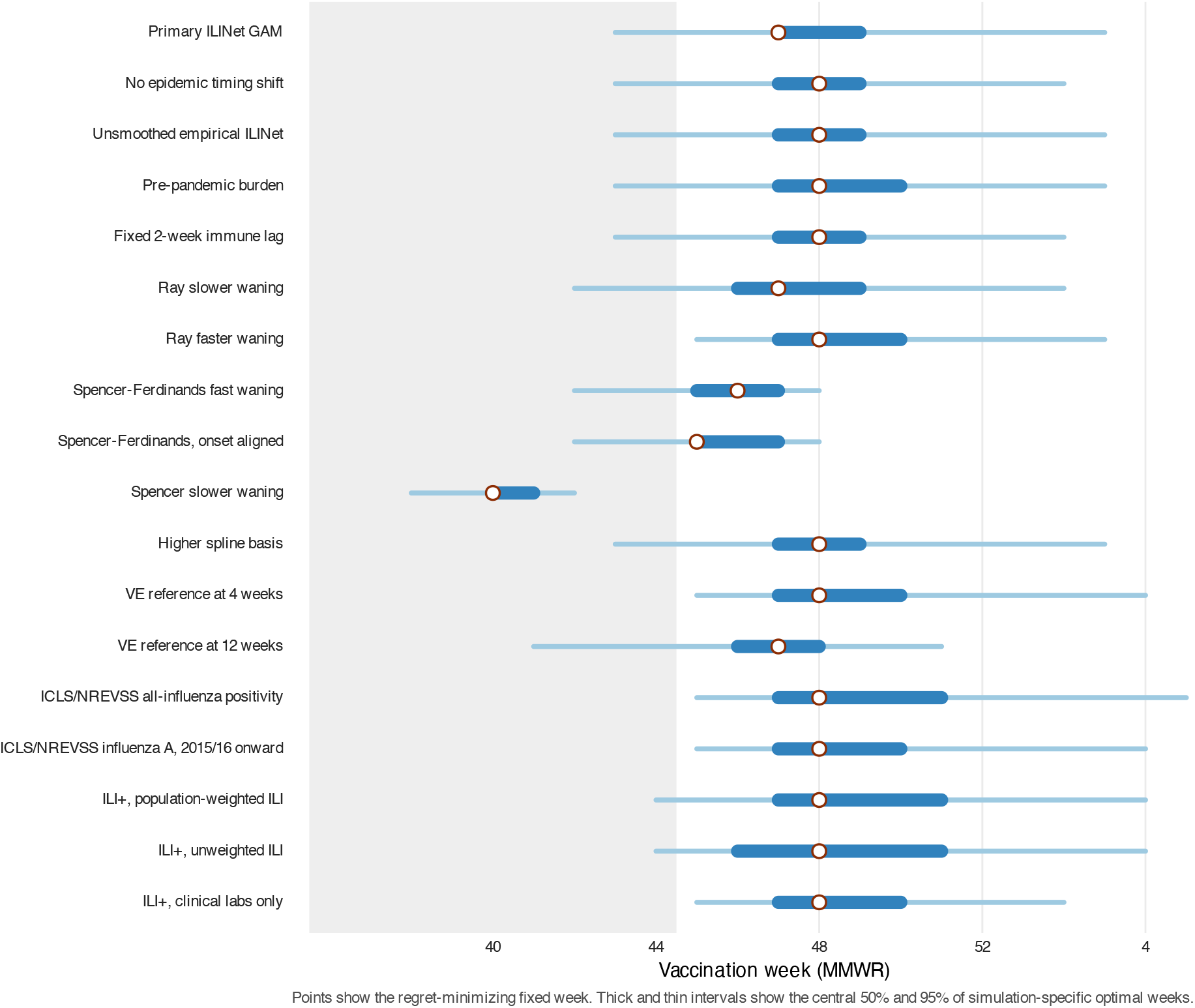
Robustness of national vaccination timing. Open points show the fixed week that minimized mean regret. Thick and thin intervals show the central 50% and 95% of best weeks across simulations. The shaded area approximates the ACIP September–October guidance window. ILI+ multiplies the national outpatient ILI proportion by national laboratory percent positivity. The clinical-laboratory-only ILI+ analysis used the six seasons with complete week 36–22 coverage from a single laboratory series.

Alternative data and smoothing choices did not move the decision back into the September– October guidance window. The unsmoothed empirical ILINet analysis and the GAM analyses with no added timing shift or more flexible fits all selected week 48. Supplement Figure S1 compares the observed curves with the primary and more flexible fits for the two seasons with visually prominent secondary peaks. Restricting the burden data to pre-pandemic seasons selected week 48.

The selected week varied with the waning model. Fixing the immune-response lag at two weeks or assuming faster Ray waning selected week 48. Slower Ray waning selected week 47. The Spencer– Ferdinands fast cubic curve selected week 46, and Spencer’s hypothetical slow cubic curve selected week 40. Vaccination in week 48 lost 2.3% of achievable modeled protection under the fast cubic curve and 9.9% under the slow curve. Changing the time at which published vaccine-effectiveness estimates were treated as fully established (4 or 12 weeks instead of 8) selected weeks 48 and 47, respectively.

Because the Spencer–Ferdinands fast cubic curve is indexed to symptom onset, whereas ILINet records the outpatient visit week, we shifted the ILINet curves earlier to approximate an onset-based timescale. The selected week moved from 46 to 45. The median best week was 46 in both analyses, and the chance that the best week had occurred by week 44 rose from 13.3% to 20.4%.

Several additional influenza-specific outcomes pointed in the same direction as the primary ILINet analysis. Analyses of national influenza percent positivity from the U.S. Influenza Collaborating Laboratories System and National Respiratory and Enteric Virus Surveillance System (ICLS/NREVSS) selected week 48, just one week later than the primary result. To ask whether non-influenza illness in syndromic surveillance was pulling the decision earlier, we repeated the analysis after weighting ILI by laboratory influenza positivity (ILI+). That influenza-weighted outpatient measure selected week 48 rather than week 47. Unweighted ILI+ also selected week 48. An ILI+ analysis restricted to the six seasons with complete weekly ILI and clinical-laboratory influenza data selected week 48. Supplement Figure S2 shows the relative-regret curves for the empirical ILINet, national ICLS/NREVSS, and main ILI+ analyses on a common scale. In the main ILI+ analysis, mean regret was 80.4 influenza-weighted outpatient visits per 10,000 weekly encounters at week 44 and 30.0 at week 48. Choosing week 48 rather than week 44 therefore reduced modeled influenza-weighted outpatient burden by about 50 visits per 10,000 weekly encounters.

Our results were also stable across seasons. When each observed season was evaluated as the burden curve on its own, the regret-minimizing week ranged from 45 to 49. In separate leave-one-season-out refits, the models trained on the other 11 seasons selected week 47 in 7 analyses and week 48 in 5. Applying each training-derived week to the omitted season cost a median of 65.0 ILI visits per 10,000 weekly encounters over that season, similar to the expectation from the primary analysis. Additional season-held-out results are provided in the Supplement.

## Discussion

For most people who will receive one influenza vaccine dose, our primary model favors vaccinating in MMWR weeks 47–48, approximately late November through early December. The primary analysis selected week 47. Week 48 performed nearly as well, and the influenza-weighted outpatient analysis selected week 48. By contrast, our analysis of children 0–4 years of age selected an earlier optimal week. State variation was modest for adults and older children, suggesting that limited geographic variation supports a common national timing estimate.

This week-level answer fills a practical gap. Current guidance recommends September or October vaccination for most people but does not explicitly quantify the early-versus-late tradeoff at the scale of an individual patient (4). While prior work showed that epidemic timing, waning, and initial protection matter (12–16), our contribution is a fixed-week estimate of optimal vaccine timing that integrates uncertainty about those factors using information available before the season. Moreover, we show that live forecasts are not required to make a good decision regarding individual vaccine timing.

It is important to note that weeks 47 and 48 should be read as a window for optimal vaccination timing rather than a reason to wait indefinitely. These weeks retained at least 95% of the protection available from the best timing in 83–86% of simulations, while week 44 did so less than half the time. Relative to the best timing, vaccination near the end of October left about 9% of achievable modeled protection against outpatient ILI surveillance burden unused, and vaccination in early October, near the midpoint of the current September–October guidance window, left about 22% unused, compared with about 2–3% in weeks 47–48. That is, vaccination in week 48 provides 19.6 percentage points more of this modeled protection than vaccination in week 40. However, if one waits an additional 4 weeks after the optimal window to get vaccinated, they will be worse off than if they had been vaccinated 4 weeks too early (within the current guidance window).

Our result is also not a claim that vaccination campaigns should wait until November. ACIP timing guidance currently appears to weigh coverage, early-season risk, and the chance that a deferred visit never happens. Our model answers a different question: given that a person will receive one dose, which week maximizes expected direct protection against outpatient ILI surveillance burden? It holds the epidemic curve and the vaccination behavior of the rest of the population fixed. A population-wide shift in timing could change vaccine coverage and influenza transmission. Our model does not estimate those population-level effects. If waiting makes later receipt uncertain, we should vaccinate when the dose can reliably be given. When a later provider visit is a good option or a patient can pick their timing through a local pharmacy, our primary model favors late November through early December for most one-dose recipients.

The earlier optimal timing result for ages 0–4 years also needs separate interpretation. Importantly, our analysis evaluates one dose and does not apply to children who need two doses that season, including many in the 0–4 years cohort and at least some people 5–8 years of age. In addition, the surveillance group includes infants younger than 6 months, who cannot be vaccinated. Furthermore, the age-specific burden values were reconstructed from regional ILI shares without age-specific outpatient denominators, and an age-specific influenza-weighted analysis was not available, so non-influenza respiratory illness could pull the pediatric curve earlier. That is because ILI syndromic surveillance captures respiratory syncytial virus (RSV) (25), and recent US data show that RSV epidemics frequently peak before influenza epidemics (26). If the ILI curves in younger demographics include a larger proportion of RSV, it may push the ILI curve earlier in that cohort and the model might select an earlier-than-optimal time of vaccination as a result. Thus, future work to better identify the optimal start of the two-dose pediatric vaccination series and data to target different age groups more specifically are needed.

The national timing estimate was stable across alternative surveillance measures and season omissions. Laboratory positivity and ILI+ selected week 48, which argues against non-influenza illness in syndromic ILI driving the late-November estimate. The waning model had a greater influence. Both confidence-limit sensitivities for Ray waning selected weeks 47–48. The Spencer– Ferdinands fast cubic curve selected week 46, with little loss of achievable protection at week 48, whereas Spencer’s hypothetical slow curve selected week 40 (15, 16). The slow curve retained protection through much of the season, reducing the benefit of delaying vaccination.

In addition, several other assumptions could still move the optimal week. Vaccine effectiveness was sampled independently of epidemic season. Correlation among subtype, effectiveness, and timing could change this balance to some extent. The primary waning estimate was used across ages and seasons (6). The model weighted vaccine protection by the temporal distribution of outpatient surveillance burden; optimal timing could differ for hospitalization or death. However, utilizing hospitalizations would produce a significant timing alignment issue in the model because protection should be evaluated at infection onset rather than outcome. That is because the influenza vaccine presumably only protects against hospitalization or death insofar as it prevents or at least attenuates influenza infection.

For a person who plans to receive one influenza vaccine dose, the primary model favored late November through early December across varied historical epidemic timing. If future work finds more sustained vaccine protection, it would likely favor earlier vaccination.

## Data Availability

All data is available through CDC. We have provided a snapshot of our development repository on github for the last pipeline run before submission. The pipeline will pull all of the data from public sources.

https://github.com/ausmeyer/flu_optimal_vaccine_estimate

## Data Availability

The ILINet and ICLS/NREVSS surveillance data analyzed in this study are publicly available through CDC FluView (https://www.cdc.gov/fluview/) and were retrieved from its current public download endpoints by code in the study repository. State population estimates are publicly available from the US Census Bureau (https://www.census.gov/data/datasets/time-series/demo/popest/2020s-state-total.html).

## Reproducible Research Statement

Code and software requirements for the complete reproducibility workflow will be publicly available at https://github.com/ausmeyer/flu_optimal_vaccine_estimate. The workflow retrieves the public source data, applies the prespecified season and MMWR-week restrictions, and regenerates the primary and sensitivity analyses, validation results, tables, and figures. Downloaded data, fitted models, and generated outputs are not stored in the repository. Each run records input hashes, analysis settings, and the software environment. Because public surveillance records and compatible software versions can change, future runs may not reproduce the reported values exactly.

## Use of Generative Artificial Intelligence

Cursor, several models, and OpenAI Codex were used to assist with reviewing and debugging code, editing manuscript text, and checking consistency across manuscript materials. The authors remain responsible for the study design, analytical decisions, code, results, interpretations, and final text.

## Author Contributions

A.G.M.: Conceptualization, Methodology, Software, Validation, Formal analysis, Investigation, Data curation, Visualization, Writing – original draft, Writing – review and editing, Project administration. B.N.R.: Conceptualization, Writing – review and editing. S.Y.: Methodology, Visualization, Writing – review and editing. M.S.: Methodology, Visualization, Writing – review and editing. All authors reviewed and approved the final manuscript.

## Disclosures

The authors declare no competing interests.

## Notes

### Competing Interest Statement

The authors have declared no competing interest.

### Author Declarations

Public ILINet and NREVSS available through CDC.

