## supplement for "Decision Analysis Modeling Favors Optimal Influenza Vaccination in Late November or Early December"

### Contents

**1 Decision Model** **3**

**2 Surveillance Data and Included Seasons** **3**

**3 Epidemic-Curve Model and Simulation** **4**

**4 Estimating Burden by Age Group** **6**

**5 Vaccine Protection** **6**

**6 Optimal Weeks, Regret, and Standardized Visits** **10**

**7 Sensitivity Analyses and Evaluation in Omitted Seasons** **11**

**8 Software and Reproducibility** **14**

**9 Supplementary Results** **14**

**References** **17**

### 1 Decision Model

We asked which Morbidity and Mortality Weekly Report (MMWR) week maximizes expected direct vaccine protection for a person who will receive one influenza vaccine dose during the season. Candidate vaccination weeks were MMWR weeks 36–52 and 1–12. We evaluated protection against surveillance burden, measured as weekly influenza-like illness (ILI) or influenza activity, from week 36 through week 22 of the following year and included week 53 when it occurred.

Each simulation draw followed the same workflow. We sampled an eligible historical influenza season and a plausible epidemic curve for that season. We then sampled baseline vaccine effectiveness (VE), the time required to develop protection, and the rate at which protection waned. For every candidate vaccination week, we multiplied the weekly epidemic curve by the protection remaining in that week and summed across the season. The candidate with the greatest total protection was the optimal week for that draw. Repeating this process produced a distribution of optimal weeks and showed how much protection was forgone (regret) when vaccination occurred earlier or later.

The analysis assumes that the person will receive one dose. It estimates direct protection for that person while holding the epidemic curve fixed. It does not account for missed vaccination caused by delay, how many people get vaccinated, or how vaccination changes influenza transmission and epidemic timing.

Table S2 summarizes the empirical inputs, published waning scenarios, and key author-specified assumptions. Figure S4 shows the resulting vaccine-protection distributions for all ages and each modeled age group.

### 2 Surveillance Data and Included Seasons

We obtained data from the U.S. Outpatient Influenza-like Illness Surveillance Network (ILINet) through the Centers for Disease Control and Prevention (CDC) FluView public data downloads on September 5, 2026 (1). Each state or jurisdiction provides the weekly number of outpatient visits for ILI and the total number of outpatient visits. ILINet collects syndromic ILI without laboratory confirmation. CDC notes that ILI can reflect any respiratory pathogen that produces fever with cough or sore throat and that surveillance data do not directly estimate the number of influenza illnesses (1).

We restricted the analytic set to the 50 states and the District of Columbia, excluding Puerto Rico and the U.S. Virgin Islands. We combined New York City and New York State records before model fitting. We began with 2010/11, the first season after the 2009 H1N1 pandemic and the first under universal U.S. influenza vaccination guidance (2). The primary season set continued through 2018/19 and then included 2023/24 through 2025/26. We excluded 2019/20 through 2022/23 to avoid seasons whose epidemic patterns were distorted by the coronavirus disease 2019 (COVID-19) pandemic, including unusually low influenza circulation and the early post-pandemic influenza pattern. The processed state, Department of Health and Human Services (HHS) regional, and national files ended on May 31, 2026, at MMWR week 22. All 12 included seasons extended through week 22 and were available for sampling in the decision model.

The burden window ran from week 36 through week 22 (roughly September through May) and covered the candidate vaccination period, immune-response lag, and late-season activity. State-level ILINet reporting began at week 40 in 2010/11. That season contributed state burden from week 40 onward, without extrapolation into the absent September weeks. The HHS, national ILINet, and national laboratory series covered weeks 36–22 in all included seasons.

Weeks with more reported outpatient visits received more weight when fitting each state's ILI curve. Weeks with no reported outpatient denominator received zero weight during fitting and were omitted from observed population-weighted summaries. We retained those weeks when evaluating the fitted curves and used the model's predictions for them. We obtained July 1, 2025 state population estimates from the United States Census Bureau to combine state results into national summaries (3).

### 3 Epidemic-Curve Model and Simulation

We placed MMWR weeks on one continuous influenza-season time axis beginning at week 36. Week 52 was followed by week 1 in most seasons and by week 53 and then week 1 when week 53 occurred. Week 53 contributed to surveillance burden but was not a candidate vaccination week.

We fit a separate penalized generalized additive model (GAM) for each state. For state $s$, season $r$, and week $t$, the response was the reported ILI proportion, $Y_{srt}/N_{srt}$, where $Y$ is the number of ILI visits and $N$ is the number of total outpatient visits. We used a quasibinomial mean-variance relationship with state-specific dispersion $\phi_s$, so that $\mathrm{Var}(Y_{srt}/N_{srt}) = \phi_s p_{srt}(1 - p_{srt})/N_{srt}$. This model gives more weight to weeks with more outpatient visits and allows greater variation than a binomial model. We did not treat visits aggregated across providers as independent individual observations.

The state model was

$$\mathrm{logit}(p_{srt}) = \alpha_s + \gamma_{sr} + f_s\{g(t)\} + b_{sr}\{g(t)\}. \quad (1)$$

Here, $p_{srt}$ is the underlying ILI proportion, $\alpha_s$ is a state intercept, $\gamma_{sr}$ is a season effect, $f_s$ is the state's shared seasonal curve, and $b_{sr}$ allows each season to depart smoothly from that curve. The function $g(t)$ gives the continuous position of week $t$ within the influenza season. We used `mgcv::gam` with a cubic regression spline of basis dimension 14 for the shared curve and a factor smooth of basis dimension 12 for the season-specific deviations; these basis dimensions set the maximum flexibility of each smooth. We estimated smoothing parameters by restricted maximum likelihood and used the default penalty multiplier $\gamma = 1$ (4).

Each state's model uses information from all included seasons but does not share coefficients with other states. We recorded deviance explained and the estimated scale parameter. We used basis-dimension checks to assess whether either smooth term was too rigid and examined residual autocorrelation at lags one through four to assess whether week-to-week correlation remained after smoothing. The model does not explicitly account for this residual correlation. We evaluated more flexible curves in a sensitivity analysis with basis dimensions 16 and 14.

Primary state fits captured a median of 97.1% of the weekly variation in ILI (minimum, 93.1%). Median within-season lag-1 residual autocorrelation was 0.023. Alaska and Kentucky were the only states in which more than one-quarter of seasons had absolute lag-1 residual autocorrelation above 0.3. Basis-dimension checks flagged 37 of 51 fits. Increasing the shared and season-specific bases to 16 and 14 reduced that number to 21 and selected week 48 in the national analysis. The unsmoothed empirical ILINet sensitivity, which does not use fitted GAM curves, also selected week 48.

Figure S1 compares the observed curves with the primary and higher-basis fits for 2018/19 and 2024/25, the two seasons with visually prominent secondary peaks. Both fits smooth some of this within-season structure. The higher-basis sensitivity selected week 48, one week later than the primary analysis.

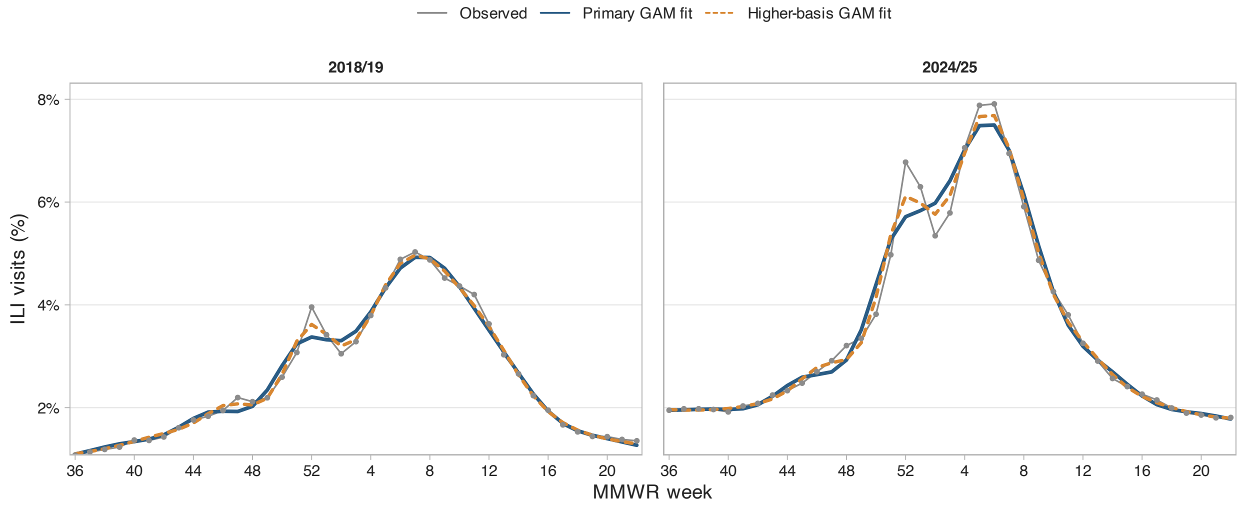


Figure S1: Observed and GAM-fitted national ILI curves for seasons with visually prominent secondary peaks. Points and thin gray lines show the observed weekly population-weighted national ILI proportion. Blue lines show the primary GAM fit, with shared and season-specific basis dimensions of 14 and 12. Orange lines show the higher-basis fit, with corresponding dimensions of 16 and 14.

To account for uncertainty in each fitted curve, we drew GAM coefficients from a multivariate normal distribution centered on the fitted coefficients with the covariance matrix returned by `mgcv`. We generated the draws with `MASS::mvrnorm` (5). These approximate empirical-Bayes draws reflect uncertainty in the fitted curve while holding the estimated smoothing parameters fixed. They are not draws from a full joint Bayesian model. We transformed each draw through the inverse-logit function to obtain a plausible weekly ILI curve.

Each simulation sampled one of the 12 eligible seasons with replacement and used that same season in every state. This preserved the observed geographic pattern of early and late seasons. We also shifted the sampled curve in time by a draw from $N(0, 0.75^2)$ weeks. We applied the same shift to every state, used linear interpolation between weeks, and held the curve's first and last values constant outside the observed range. This shift allows simulated epidemic timing to vary around the observed historical seasons. We specified its standard deviation as a modeling assumption.

For national summaries, we averaged the state curves using July 1, 2025 state population weights. This follows CDC's approach to national ILI summaries and prevents states from contributing in proportion to ILINet reporting volume (1). The resulting state and national curves remain smoothed syndromic ILI proportions.

Figure S3 compares the normalized national seasonal curves for primary ILI, laboratory influenza positivity, and population-weighted ILI+. Normalization is used only to display seasonal timing on a common scale. The decision analyses retain each curve's burden scale.

### 4 Estimating Burden by Age Group

The public state-level ILINet extract contains all-age state counts. It does not contain the age-specific outpatient denominators needed to fit state-age ILI proportions. We therefore used a two-stage model. The first stage generated the all-age state curve. The second stage used the distribution of ILI visits within each HHS region to divide that curve among the five ILINet age groups, following the geographic allocation used by Spencer and colleagues (6).

For each sampled season, HHS region, and week, we drew the five age shares from a Dirichlet distribution with parameters equal to the reported age-specific ILI visit counts plus 0.5. The one-half count provides light smoothing for small or zero cells. We multiplied each state's all-age ILI curve by the sampled age share from its HHS region. For age-specific analyses, we applied the timing shift after this multiplication.

Age-specific timing depends on weekly changes in the regional age distribution of ILI visits and on age-matched VE. The model does not construct an age-specific outpatient denominator. It assumes that the HHS-region age distribution of ILI visits represents each state in that region.

### 5 Vaccine Protection

We assembled adjusted test-negative VE estimates for medically attended influenza from CDC tables and the source literature. The 2010/11 estimates came from Treanor and colleagues, and the 2011/12 through 2018/19 estimates came from archived CDC tables (7, 8). For 2015/16 overall VE, we used the confidence interval in the primary report rather than the conflicting interval on the archived CDC page (9). We added final overall and age-specific estimates for 2023/24 (10), the final overall U.S. Flu VE Network outpatient estimate for 2024/25 (11), and the preliminary overall estimate for 2025/26 (12). Thus, the all-age pool contained 12 season-specific estimates and each age-specific pool contained 10. Table S1 lists every estimate, confidence interval, and source.

We matched model age groups to the closest age ranges available in the VE studies. Ages 0–4 years used estimates for ages 6 months–8 years, except that the 2023/24 source group began at 8 months. Ages 5–24 years used estimates for ages 9–17 years as a proxy. Ages 25–49 years used estimates for ages 18–49 years. The 2010/11 estimate for ages 9–49 years supplied both of these pools. Ages 50–64 and 65+ years used estimates for matching age groups. The 0–4 ILI group includes infants who are too young to receive influenza vaccine. The VE proxy for ages 5–24 years does not directly represent adults aged 18–24 years in most source seasons.

For each draw, we sampled one season-specific VE estimate with replacement from the relevant all-age or age-matched pool, giving each source season equal probability independently of the epidemic season. Because test-negative VE equals one minus an adjusted odds ratio, we converted each VE estimate and its confidence interval to the log odds-ratio scale. We sampled a log odds ratio from the resulting normal distribution and exponentiated it to obtain a reported effect ratio.

Published seasonal VE estimates average outcomes observed at different times since vaccination and may already reflect waning during the season. We treated each sampled estimate as VE eight weeks after full immune response and used the sampled waning rate to estimate VE at the start of full protection. We chose this reference time because the source tables do not report one common time since vaccination. After drawing a waning rate $\beta$, we divided the reported effect ratio by $\exp(8\beta/4)$ and converted the result back to baseline VE. We bounded baseline VE to $[0, 1]$, recorded when this limit was applied, and examined reference times of 4 and 12 weeks in sensitivity analyses.

CDC states that immune protection develops about two weeks after vaccination (2). We allowed full immune response to occur after one, two, or three weeks with probabilities 0.15, 0.70, and 0.15. CDC guidance supports the two-week center of this distribution. The probability weights are author-specified.

For waning, Ray and colleagues estimated that the odds of influenza increased by a factor of 1.16 for each additional 28 days since vaccination, with a 95% confidence interval of 1.13 to 1.20 (13). We sampled the log odds-ratio multiplier $\beta$ from a normal distribution with mean log(1.16) and standard error reconstructed from those confidence limits. If vaccination occurred in week $w$, we calculated the number of weeks $H_{wt}$ between full immune response and surveillance week $t$. Protection was zero before full immune response. Afterward, protection in draw $d$ and age group $a$ was

$$q_{awt}^{(d)} = 1 - \left(1 - VE_{0a}^{(d)}\right) \exp\left\{\beta^{(d)} H_{wt}^{(d)}/4\right\}. \tag{2}$$

We bounded protection to $[0, 1]$. Dividing by 4 converts weeks to 28-day periods. We assumed that the waning association reported by Ray and colleagues applies to every modeled season and age group, with the waning multiplier acting on the baseline effect ratio. The curve describes declining protection without modeling the underlying immune mechanisms.

A reanalysis of the same data restricted to people vaccinated before influenza circulation found a similar odds ratio of 1.18 per 28 days (95% confidence interval, 1.12–1.23) (14).

Table S1: Vaccine-effectiveness estimates used in the simulations. Values are reported effectiveness percentages and 95% confidence intervals before uncertainty sampling and calibration to the onset of full immune response.

| Season | All ages | 6 months–8 years | 9–17 years | 18–49 years | 50–64 years | ≥65 years | Source |
|---|---|---|---|---|---|---|---|
| 2010/11 | 60 (54, 66) | 63 (52, 72) | 51 (36, 62) | 51 (36, 62) | 51 (25, 68) | 36 (−22, 66) | (7) |
| 2011/12 | 47 (36, 56) | 45 (20, 62) | 58 (27, 76) | 44 (21, 60) | 54 (23, 72) | 43 (−18, 72) | (15) |
| 2012/13 | 49 (43, 55) | 57 (45, 67) | 39 (18, 54) | 39 (26, 50) | 65 (54, 74) | 26 (−10, 50) | (16) |
| 2013/14 | 52 (44, 59) | 45 (20, 63) | 53 (23, 72) | 54 (41, 64) | 59 (43, 70) | 50 (16, 71) | (17) |
| 2014/15 | 19 (10, 27) | 25 (6, 40) | 25 (2, 42) | 7 (−12, 33) | 20 (−3, 38) | 32 (3, 52) | (18) |
| 2015/16 | 48 (41, 55) | 51 (33, 64) | 59 (36, 74) | 52 (39, 61) | 26 (2, 44) | 42 (6, 64) | (9, 19) |
| 2016/17 | 40 (32, 46) | 57 (43, 68) | 36 (15, 52) | 19 (0, 34) | 40 (24, 53) | 20 (−11, 43) | (20) |
| 2017/18 | 38 (31, 43) | 68 (55, 77) | 32 (16, 44) | 33 (21, 44) | 30 (13, 44) | 17 (−14, 39) | (21) |
| 2018/19 | 29 (21, 35) | 48 (37, 58) | 7 (−20, 28) | 25 (10, 37) | 14 (−10, 33) | 12 (−31, 40) | (22) |
| 2023/24 | 44 (36, 51) | 68 (51, 79) | 59 (35, 75) | 38 (24, 50) | 16 (−11, 41) | 37 (5, 58) | (10) |
| 2024/25 | 33 (24, 41) | – | – | – | – | – | (11) |
| 2025/26 | 24 (8, 38) | – | – | – | – | – | (12) |

The all-age analyses sampled 12 season-specific estimates; each age-specific analysis sampled 10. Dashes indicate estimates not used in that pool. All rows concern medically attended influenza A or B. The 2024/25 row is a final U.S. Flu VE Network outpatient estimate; the 2025/26 row is preliminary.

Age headings identify the VE pools. Model ages 0–4, 5–24, and 25–49 years use the first three age-specific pools, respectively; model ages 50–64 and ≥65 years use matching pools. For 2010/11, the 6 months–8 years estimate comes from the combined-age result in the article text, and the published 9–49 years estimate supplies both the 9–17 and 18–49 years pools. For 2023/24, the youngest source group is 8 months–8 years. The recent all-age estimates also begin at age 8 months.

For 2015/16, the overall confidence interval is taken from Jackson and colleagues (41–55%); the archived CDC page gives a conflicting interval of 43–55%. The age-specific values for that season come from the archived CDC table. The companion CSV records the exact source URL, source age group, table location, and publication status for every estimate.

Table S2: Empirical inputs, published waning scenarios, and key parameter distributions. Table S1 lists individual VE estimates and their sources.

| Input | Value or distribution | Source and adaptation |
|---|---|---|
| **External inputs and published scenarios** | | |
| Vaccine effectiveness (VE) | 12 all-age estimates and 10 per age-specific pool, with 95% confidence intervals (Table S1). Sample source seasons equally and independently of epidemic season; sample normal log odds ratios using each estimate and interval. | Cited in the VE input table; sampling choices are ours. |
| Time to full immune response | About 2 weeks. Use 1, 2, or 3 weeks with probabilities 0.15, 0.70, and 0.15; fixed 2 weeks in sensitivity analysis. | CDC supports the 2-week center; distribution and sensitivity are our choices. (2) |
| Primary waning | Odds ratio 1.16 (95% CI, 1.13–1.20) per 28 days. $\beta \sim N\{\log(1.16), s^2\}$, where $s = \dfrac{\log(1.20) - \log(1.13)}{2\Phi^{-1}(0.975)}$. Fixed endpoint sensitivities use $\beta = \log(1.13)$ or $\log(1.20)$. | Ray et al.; normal reconstruction and fixed-endpoint sensitivities are our choices. (13) |
| Alternative waning scenarios | For $H$ weeks after full response, let $b = H/2$: Fast $r(H) = \left[\dfrac{55 - 1.37b + 0.18b^2 - 0.03b^3}{55}\right]_0^1$. Slow $r(H) = \left[\dfrac{55 - 0.50b + 0.05b^2 - 0.01b^3}{55}\right]_0^1$. The cubic coefficients use two-week units; division by 55 gives relative retention. | Spencer et al. fast and slow scenarios; the fast curve draws on Ferdinands et al., whereas the slow curve is hypothetical. (6, 23) |
| Onset-to-enrollment delay | 0-2 days: 8,520; 3-4 days: 10,754; 5-7 days: 7,861 (n=27,135). For onset alignment, sample categories in proportion to these counts and an integer day uniformly within the selected category. Shift burden earlier by that delay; primary delay is zero. | Balasubramani et al. counts summed across sites; within-category distribution and alignment are our choices. (24) |
| National population weights | July 1, 2025 populations for 51 jurisdictions (50 states and DC), held fixed across seasons and age groups. | U.S. Census Bureau; fixed-year weighting is our choice. (3) |
| **Key author-specified assumptions** | | |
| VE reference time | 8 weeks after full response; 4 and 12 weeks in sensitivity analyses. Estimate initial VE using VE at the reference time and the sampled waning curve. | Modeling assumption; no common reference time is reported by the source studies. |
| Additional epidemic timing shift | $N(0, 0.75^2)$ weeks, shared across states; SD=0 in the no-shift sensitivity. | Modeling assumption. |

$[x]_0^1 = \min\{\max(x, 0), 1\}$; $\Phi^{-1}$ is the standard-normal quantile. CDC indicates Centers for Disease Control and Prevention; CI, confidence interval; DC, District of Columbia; SD, standard deviation. Model fitting, protection equations, and other analysis settings are described in the Methods and reproducible code.

#### 6 Optimal Weeks, Regret, and Standardized Visits

For each draw, state $s$, age group $a$, and candidate vaccination week $w$, we calculated utility as

$$U_{saw}^{(d)} = \sum_{t \in \mathcal{T}} B_{sat}^{(d)} q_{awt}^{(d)}, \tag{3}$$

where $B_{sat}^{(d)}$ is the sampled ILI burden and $q_{awt}^{(d)}$ is modeled vaccine protection. The sum covers week 36 through week 22. Utility is the amount of ILI surveillance burden covered by vaccine protection.

The optimal week in each draw was the candidate with the greatest utility. We selected randomly among numerical ties using a relative tolerance of $10^{-9}$ and retained the number of tied maxima. Across 5,000 draws, we summarized the probability that each week was optimal and calculated the median and central 50% and 95% intervals. We calculated these summaries on the continuous influenza-season axis before converting them back to MMWR week labels. We also reported the cumulative probability that the optimal week had occurred by exact MMWR weeks 44, 48, and 52.

Simulation error at 5,000 draws is small relative to the differences we report. The 95% Monte Carlo half-width was at most 1.4 percentage points for the primary cumulative probabilities and the near-optimality probabilities. Probabilities resting on one or two draws are reported as thresholds rather than point values.

Regret measures how much modeled protection was lost relative to the best week in the same draw:

$$U_{sa}^{(d),\text{best}} = \max_{w} U_{saw}^{(d)}, \tag{4}$$

$$R_{saw}^{(d)} = U_{sa}^{(d),\text{best}} - U_{saw}^{(d)}. \tag{5}$$

We averaged regret across draws for each candidate week. The week with the lowest mean regret is also the week with the highest mean utility across draws.

For clinical presentation, we multiplied ILINet regret and its interval limits by 10,000. This expresses the modeled ILI burden left avoidable relative to the best week in each simulation as standardized visits over the season in a system with 10,000 outpatient encounters each week. This change of scale preserves the estimated optimal weeks and comparisons among candidate weeks. The best week can differ across simulations, so mean regret can remain above zero even for the best fixed vaccination week.

We divided regret by the best utility in each draw to calculate relative regret. We treated a candidate week as near optimal when its utility was within 5% of the best utility for that draw. If best utility was zero, we set relative regret to zero for every week.

### 7 Sensitivity Analyses and Evaluation in Omitted Seasons

We repeated the all-age decision analysis under alternative assumptions. Each sensitivity used 5,000 draws and retained the primary settings unless stated below.

To assess the added epidemic timing shift, we repeated the primary GAM analysis with its standard deviation set to zero. We retained the same fitted curves, sampled seasons, coefficient draws, and vaccine-protection draws.

For the empirical ILINet sensitivity, we sampled eligible national ILINet seasons that extended through week 22 with replacement, using the observed data without fitting GAM curves. For each week, we drew the unweighted ILI proportion from a beta distribution with shape parameters $Y + 0.5$ and $N - Y + 0.5$, multiplied it by the observed ratio of CDC's population-weighted to unweighted ILI percentages, and bounded the result to $[0, 1]$. We applied the same timing shift used in the primary analysis directly to these sampled weekly curves. The shift could include a fraction of a week, so we used linear interpolation between weekly values and held the first and last values constant beyond the observed range.

To evaluate the influence of the three recent burden seasons, we restricted the burden season set to 2010/11 through 2018/19 while retaining the expanded all-age VE pool.

In separate analyses of vaccine-protection assumptions, we fixed the immune-response lag at two weeks or fixed the Ray waning odds ratio at the lower or upper limit of its reported 95% confidence interval. We also used Spencer and colleagues' fast cubic waning curve, adapted from Ferdinands and colleagues, and their hypothetical slow cubic curve (6, 23). For $H$ weeks after full immune response, we set $b = H/2$ to express elapsed time in two-week units. The fractions of initial VE remaining were

$$r_{\text{fast}}(H) = \frac{55 - 1.37b + 0.18b^2 - 0.03b^3}{55}, \tag{6}$$

$$r_{\text{slow}}(H) = \frac{55 - 0.50b + 0.05b^2 - 0.01b^3}{55}. \tag{7}$$

We bounded both retention curves to $[0, 1]$. We calibrated initial VE so that each sampled source estimate applied at the primary eight-week reference time.

We assessed curve flexibility by increasing the shared and season-specific GAM basis dimensions from 14 and 12 to 16 and 14. In separate analyses, we changed the VE reference time to 4 or 12 weeks. We also fit national GAMs to weekly percent positivity from the U.S. Influenza Collaborating Laboratories System and National Respiratory and Enteric Virus Surveillance System (ICLS/NREVSS) (1). The all-influenza analysis used combined public health and clinical laboratory data before 2015/16. From 2015/16 onward, we used clinical laboratory data when available and combined data otherwise. Combined data were needed for MMWR weeks 36–39 of 2015/16. A second analysis used influenza A positivity from that 2015/16-onward series. The denominator in these analyses is tests performed, so we reported timing and relative regret without converting absolute regret to standardized visits.

To obtain a more influenza-specific outpatient measure, we constructed weekly national ILI+ as the product of the ILINet ILI proportion and the ICLS/NREVSS influenza positivity proportion. Multiplying the proportion of outpatient visits for ILI by the proportion of tested specimens positive for influenza has been used to estimate influenza-associated outpatient ILI (25). ILINet and ICLS/NREVSS are separate sentinel systems in our analysis, so ILI+ is an influenza-attributable outpatient proxy rather than a count of laboratory-confirmed visits.

For each sampled week, we drew the unweighted ILI proportion from a beta distribution with shape parameters $Y + 0.5$ and $N - Y + 0.5$. For the population-weighted analysis, we multiplied this draw by the observed ratio of the CDC population-weighted to unweighted ILI proportions and bounded the result to $[0, 1]$. We independently drew ICLS/NREVSS positivity from a beta distribution with shape parameters equal to positive tests plus 0.5 and negative tests plus 0.5, then multiplied the two sampled proportions. Within each sampled season, we fit a cubic smoothing spline to $\log(1+p)$ with smoothing parameter 0.65, transformed predictions back to the proportion scale, and applied the same timing shift used in the primary analysis.

We evaluated three ILI+ specifications. The main ILI+ analysis used population-weighted ILI and all 12 seasons. A second used unweighted ILI and the same seasons. Both used the national laboratory series described above. A third analysis used population-weighted ILI and the six seasons with a complete week 36–22 burden window from the clinical laboratory series alone: 2016/17 through 2018/19 and 2023/24 through 2025/26. We multiplied absolute regret by 10,000 and report the result as ILI+-weighted outpatient visit equivalents in a surveillance system with 10,000 encounters each week. Differences between candidate weeks represent the additional visit equivalents potentially averted by changing vaccination timing. This is a standardized proxy for influenza-attributable outpatient burden.

Figure S2 compares the relative-regret curves for the empirical ILINet, national all-influenza ICLS/NREVSS, and main population-weighted ILI+ analyses. Expressing regret as a percentage of achievable protection allows comparison across these different surveillance measures.

The Spencer–Ferdinands fast-waning curve comes from an analysis that measured time since vaccination at symptom onset, whereas ILINet records the outpatient visit week. We therefore performed a paired sensitivity analysis that shifted the ILINet curve earlier to approximate symptom-onset timing. We used the time from symptom onset to enrollment reported for 27,135 United States Influenza Vaccine Effectiveness Network outpatients with complete interval data in 2011–2016. Of these, 8,520 enrolled 0–2 days after onset, 10,754 enrolled after 3–4 days, and 7,861 enrolled after 5–7 days (24). For each draw, we sampled an interval according to these counts and then selected a whole number of days within that interval, giving each day equal probability. We applied the sampled shift to every jurisdiction in the draw. We did not apply this shift to the primary Ray analysis because Ray and colleagues measured time since vaccination at the polymerase chain reaction (PCR) test date, which is close to the ILINet visit date.

We refit the model while omitting each season in turn to assess whether any single season drove the timing estimate. For each omitted season, we selected the regret-minimizing week using the

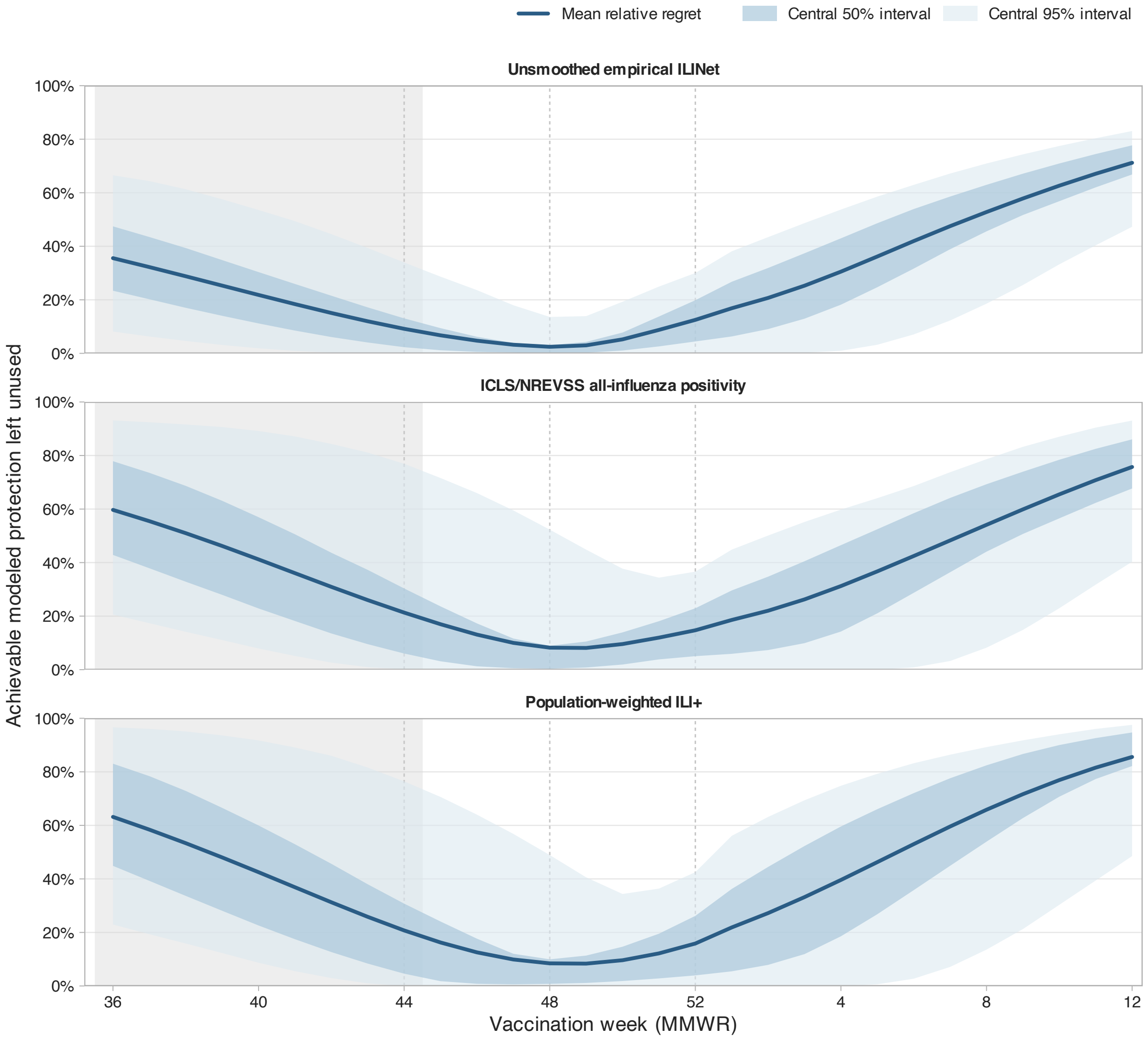


Figure S2: Relative regret by vaccination week for alternative surveillance outcomes. Relative regret is the percentage of modeled protection left unused compared with the best week in the same simulation; lower values indicate better timing. Solid lines show mean relative regret, and bands show the central 50% and 95% of simulation-specific relative regret. Because relative regret expresses loss as a fraction of achievable protection in each simulation, the week minimizing mean relative regret can differ from the week selected by mean absolute regret. Dashed lines mark weeks 44, 48, and 52; the shaded region approximates the ACIP September–October guidance window. The empirical ILINet analysis used an unsmoothed bootstrap of observed weekly ILI values rather than GAM-fitted burden draws. The ICLS/NREVSS analysis used national all-influenza percent positivity. The ILI+ analysis multiplied population-weighted ILI by influenza positivity and included all 12 seasons. All panels use the same vertical scale.

other 11 seasons. We then evaluated that week against the observed population-weighted national ILI curve in the omitted season. We held that curve fixed while sampling uncertainty in VE, waning, and immune-response lag across 5,000 draws. We recorded standardized visits lost and relative regret. We also calculated the fraction of best weeks in the omitted-season simulations that fell within the 50% and 95% intervals estimated from the other seasons.

### 8 Software and Reproducibility

We conducted the analysis with R version 4.5.1 (26). Code in the study repository retrieved surveillance records directly from the current CDC FluView public data downloads. We fit penalized GAMs with `mgcv` (4), sampled multivariate normal coefficients with `MASS` (5), and created figures with `ggplot2` (27). The scripts set random-number seeds. Saved results include the optimal week in each draw, probabilities that the best week had occurred by specified weeks, sampled VE and waning values, model diagnostics, regret curves, near-optimality curves, and sensitivity summaries.

The public repository contains the code and software requirements rather than downloaded data, fitted models, or generated results. By default, the analysis pipeline downloads the current public records, applies the prespecified season and MMWR-week restrictions, checks the resulting inputs, and regenerates every reported analysis and figure. It requires minimum package versions. Each run records the versions of R, its packages, the computing platform, and linked software libraries. Public surveillance records and compatible software versions can change, so later runs may produce values that differ from those reported here.

The scripts reuse a saved GAM only when its filename matches the included seasons, smoothing settings, latest observed MMWR week, and a checksum of the input data and fitting code. Each analysis writes a JavaScript Object Notation (JSON) file beside its tables. This file records input file checksums, the path to the saved model, and the main analysis settings. Primary results, sensitivity results, and model diagnostics are saved in separate directories.

### 9 Supplementary Results

#### 9.1 Evaluation in Omitted Seasons

The season-specific and leave-one-season-out analyses addressed complementary questions. When each observed season was used as the burden curve on its own, the regret-minimizing week ranged from 45 to 49, and the median of the 12 season-specific median optimal weeks was 49. In the leave-one-season-out evaluation, models fit to the other 11 seasons selected week 48 after omitting 2012/13 through 2014/15, 2023/24, or 2025/26 and week 47 in the other 7 analyses. Applying each selected week to the omitted season produced a median regret of 65.0 standardized ILI visits per 10,000 weekly outpatient encounters over the season. Across the 12 omitted-season evaluations, a mean of 60.7% of best weeks in the simulations fell within the central 50% interval estimated from the other seasons, and 95.1% fell within the corresponding central 95% interval. The intervals

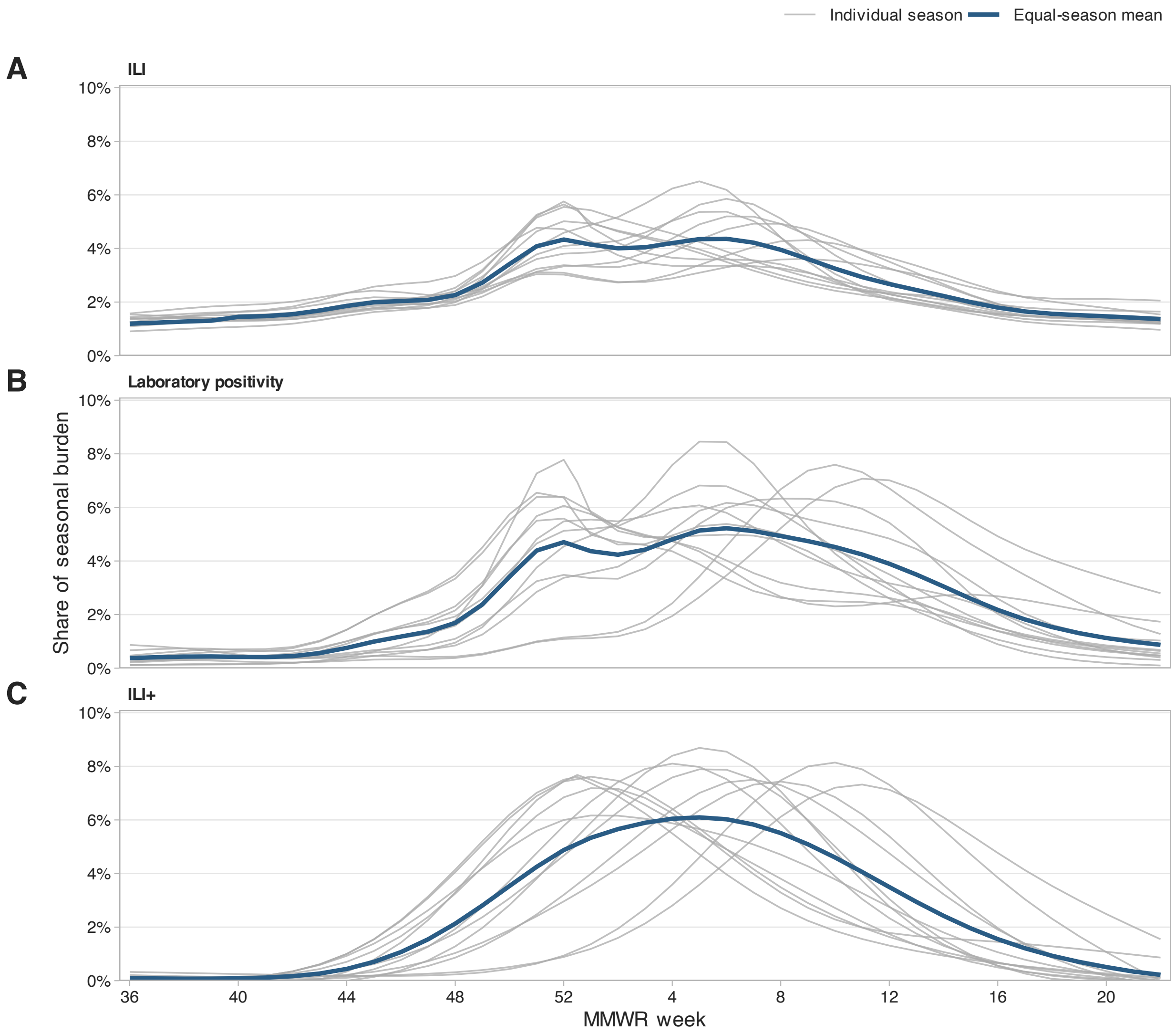


Figure S3: National surveillance curves across the 12 included seasons. Thin gray lines show each seasonal curve divided by its total burden. Blue lines show the mean with each season weighted equally. ILI is the population-weighted mean of state GAM predictions. Laboratory positivity is the national all-influenza GAM prediction. The laboratory series combines public health and clinical laboratories before 2015/16 and preferentially uses clinical laboratories thereafter. ILI+ is population-weighted national ILI multiplied by influenza positivity and smoothed with the cubic spline used in the analysis. Curves are shown before the timing shifts and do not show simulation uncertainty. Curves are aligned by MMWR week. Week 53 is shown only for individual seasons that contain it. The ILI curve derived from state data for 2010/11 begins at week 40. Its missing September weeks contribute zero to the displayed ILI mean. All other curves begin at week 36. ILI indicates influenza-like illness; GAM, generalized additive model; MMWR, Morbidity and Mortality Weekly Report.

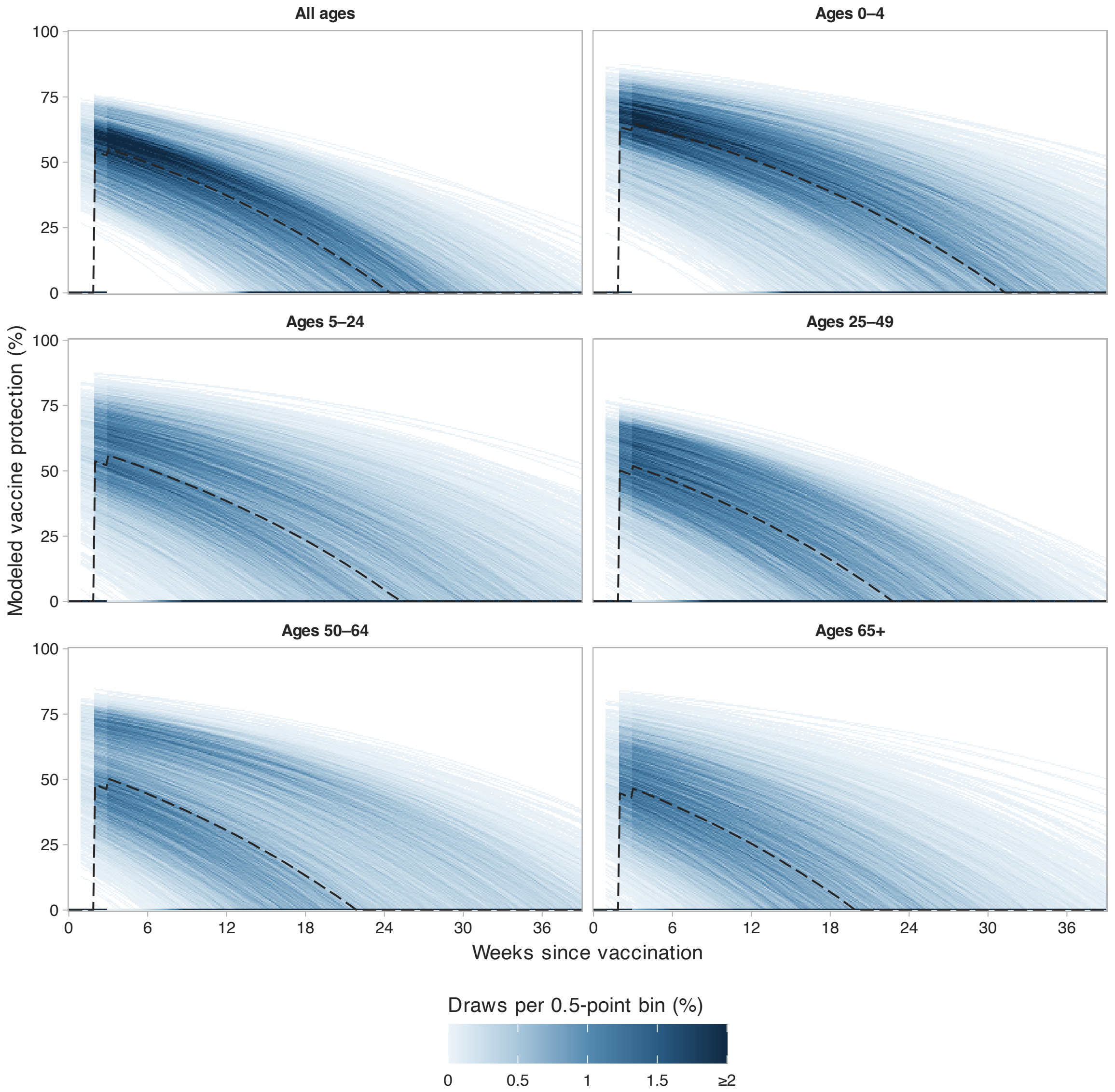


Figure S4: Distribution of modeled vaccine protection after vaccination. Each panel uses the 5,000 saved draws from the corresponding all-age or age-specific primary analysis. Shading shows the percentage of all draws in each half-percentage-point range of vaccine protection. The color scale is linear from 0 to 2%, with values of 2% or more shown in the darkest shade. Curves are evaluated daily for display. The decision analysis remains weekly. White cells contain no draws, and black dashed lines show medians. The lowest bin includes zero protection. Curves incorporate the sampled source VE, immune-response lag, and Ray waning rate. Initial VE is estimated from VE eight weeks after full response using the sampled waning rate, and protection is bounded to $[0, 1]$. Protection is zero before the sampled one-, two-, or three-week response lag. The plot shows the distribution across simulations. Tables S1 and S2 provide the inputs, distributions, and sources. VE indicates vaccine effectiveness.

covered most of the omitted-season timing distributions, and the selected week varied by only one week across omissions.
